# ACMG Secondary Findings in the Brazilian Rare Genomes Project: Insights from 5,402 genome sequencing

**DOI:** 10.1101/2025.01.22.25320957

**Authors:** Eduardo Perrone, Luiza Virmond, Antonio Victor Campos Coelho, Marina De França, Carolina Araújo Moreno, Joana Rosa Marques Prota, Jessica Grasiela de Araujo Espolaor, Michele Migliavacca, Thiago Yoshinaga Tonholo Silva, Caio Robledo D’Angioli Costa Quaio, José Ricardo Magliocco Ceroni, Kelin Chen, Renata Moldenhauer Minillo, Anne Caroline Barbosa Teixeira, Renata Yoshiko Yamada, Vivian Pedigone Cintra, Lucas Santos de Santana, Gabriela Pereira Campilongo, Renata Martins Ribeiro da Silva, Karla de Oliveira Pelegrino, João Bosco de Oliveira Filho, Tatiana Ferreira de Almeida

## Abstract

**Purpose:** Secondary findings (SF) are pathogenic or likely pathogenic variants in genes unrelated to the primary purpose of genetic testing. The American College of Medical Genetics (ACMG) provides guidelines on which SF should be reported, involving 81 genes linked to different conditions. With the increasing use of genome sequencing (GS), SF are more frequently detected, presenting challenges for healthcare systems. The Brazilian population is often underrepresented in genomic studies, which limits population-specific knowledge.

**Objective:** This study aimed to outline the profile of SF in the Brazilian Rare Genomes Project (BRGP).

**Methods:** We analyzed retrospectively SF (ACMG) data from GS of 5,402 BRGP individuals.

**Results:** Of the 5,316 cases who consented to receive SF, 3.6% (191 cases) had at least one SF. The most common genes identified were *TTR, TTN,* and *BRCA2*. SF were mainly related to cardiovascular conditions (40.2%) and cancer predisposition (37.6%). Some variants, such as *TTR:* c.424G>A; p. (Val142Ile) and *TP53:* c.1010G>A; p. (Arg337His), were recurrent, reflecting population-specific traits and founder effects. Novel variants were 10.6% of SF.

**Conclusion:** SF rate varies across studies and populations. While SF can aid early diagnosis, their relevance is debated due to potential psychological and healthcare burdens. Effective genetic counseling and public health policies are essential.

## INTRODUCTION

Secondary findings (SF), previously known as incidental findings, were defined by American College of Medical Genetics (ACMG) as a result of a deliberate search for pathogenic and likely pathogenic (P/LP) variants in genes that are not apparently relevant to a diagnostic indication for which the sequencing test was ordered.^1^ Initially, ACMG recommended to report P/LP variants in a list of 56 genes, considered actionable, despite the willingness of the individual. This gene list has been updated periodically, as new evidence of actionable genes arises. The latest list contains 81 genes.^2^

The clinical conditions associated with these genes include hereditary cancer predisposition syndromes, cardiovascular diseases, inborn errors of metabolism, among others. These conditions are primarily considered in this context due to the potential benefits of an early diagnosis.^1,2^

In the last decades, there has been a progressive increase in the use of massive parallel sequencing for clinical and research purposes. Consequently, secondary findings will become a reality in the clinical practice of healthcare professionals, who must be prepared to deal and to manage individuals with these findings. Furthermore, the healthcare system must be prepared for the health burden that SF may cause.^3,4^

Some studies have already described the percentage, and the profile of SF identified either in control or affected populations. Moreover, it is already known that some variants are typically more identified in individuals according to their genomic ancestry, so studies focusing on SF in specific populations other than Caucasian are fundamental.^5^ Brazilian population is recognized as one the most admixed in the world and to our knowledge few studies analyzed the profile of SF in a cohort of Brazilian individuals ^6,7,8^

The Brazilian Rare Genomes Project (BRGP) is an initiative of Hospital Israelita Albert Einstein (HIAE) in partnership with the Brazilian Ministry of health which aims to shorten the diagnostic journey of individuals with rare genetic diseases and create a Brazilian database of genomic variants through whole genome sequencing (WGS). Herein, our objective is to describe molecular data of SF identified in a sample of 5,402 individuals who were enrolled in BRGP.

## MATERIALS AND METHODS

### The Brazilian Rare Genomes Project and Ethical Aspects

The BRGP is a publiclllprivate initiative which aims to perform WGS in individuals with rare genetic diseases, including hereditary cancer predisposition syndromes. This project involves services from the public health system across the different regions of Brazil (South, Southeast, Midwest, North, and Northeast). There are no age restrictions and individuals of both sexes may be included in the project. Clinical data are provided by the participating centers through an online platform, using Human Phenotype Ontology (HPO) terms.

Individuals included are classified into 20 distinct clinical cohorts based on their clinical findings, which encompass: neurological disorders, connective tissue disorders, hereditary cancer syndromes, cardiological conditions, critically ill patients, dermatological diseases, skeletal dysplasias, kidney diseases, endocrinological disorders, inborn errors of metabolism, hematological diseases, inborn errors of immunity, neurocutaneous disorders, neuromuscular diseases, ophthalmological conditions, otorhinolaryngological diseases, pulmonary diseases, established genetic syndromes, gastrointestinal disorders, and vascular diseases.

A written informed consent is obtained from each participant, which includes the option to accept or decline receiving results related to secondary findings (following recommendations by ACMG). In BRGP, the reporting of such findings is only conducted in cases where consent has been provided regardless of the participant’s age. The BRGP and related studies were submitted to the ethics committees and received approval (CAAE: 9567220.4.1001.0071.).

### Genome Sequencing, bioinformatics and SF data analysis

The complete technical protocol used for WGS in BRGP is comprehensively detailed in Coelho et al.^9^ In summary, genomic DNA was extracted from peripheral whole blood and sequencing was performed on the Illumina NovaSeq 6000. Then, FASTQ files were aligned to the GRCh38/hg38 human genome reference. Variant calling was performed using DRAGEN germline pipeline from Illumina, variants were annotated using in-house protocols, according to the Matched Annotation from NCBI and EMBL-EBI (MANE) transcript^10^ and to the International System for Human Cytogenomic Nomenclature for structural variants (SV).^11^ The data analysis was performed using an online platform (Varstation®).

Only pathogenic/likely pathogenic (P/LP) variants (according to ACMG recommended criteria)^12,13^, and which are aligned with the ACMG recommendations for reporting SF^2^ are reported. It is worth noting that when the WGS analyses in BRGP began in 2020, the version of the gene list recommended by the ACMG was v2.0 ^14^ As newer versions were released, the project continuously adapted its analysis practices to include these updated gene lists.^2,14,15,16^

### Study design

In the present study we conducted a retrospective data analysis of 5,402 reports previously released by the BRGP (from December 2020 to May 2024). Individuals who had at least one ACMG secondary finding reported in their results were included. Those who did not consent to receive secondary findings were excluded.

### Statistics

Categoric variables were calculated in terms of their frequencies and continuous variable (age), in terms of their median (minimum-maximum value). A comparison between the frequency of secondary findings rate in different ages groups (≥ 60 years old versus < 60) was performed using chi-squared test. A p-value < 0.05 was considered statistically significant.

## RESULTS

### Sample Characterization

Out of the 5,402 unrelated cases initially evaluated, 1.6% (86/5,402) were excluded due to their refusal to consent to receive SF data. Among the remaining 98.4% (who provided consent) (5,316/5,402), 3.6% (191/5,316) had at least one SF variant. Out of the remaining 5,125 without secondary findings, the median age was 13 years old, varying from 0 to 92 years. Regarding different age groups, 95.7% (4905/5125) were < 60 years old and 4.3% (220/5125) were ≥ 60 years old.

### Secondary findings data

The 191 cases with SF consisted of 47.6% of males (91/191) and the median age of our sample was 11 years, varying from 0 to 77 years old. Based on the different age groups, the individuals were distributed as follows: 96.3% (184/191) were < 60 years old - the majority being < 18 years old (63.9%:122 out of 191) - and 3.7% (7/191) were ≥ 60 years. Chi-square test showed no statistical significance between the rate of secondary findings in individuals < 60 years old versus ≥ 60 years old (p value = 0.67).

Most cases came from Southeast region of Brazil (56% - 107/191), followed by Northeast (33% - 63/191), South (7.3%-14/191) and Midwest (3.7% - 7/191).

Regarding the primary findings status for these cases, 31% (59/191) had their final report with a positive result for a primary finding, while 69% (132/191) had a non-positive status.

The more represented groups of conditions related to SF were cardiovascular conditions (40.2% - 78/194), cancer predisposition syndromes (37.6% - 73/194), miscellaneous phenotypes (21.1% - 41/194) and inborn errors of metabolism (1% - 2/194) (Figure 2). Within these groups, a predominance of specific conditions was observed. In the cardiovascular group, *TTN* gene-related cardiomyopathy (MIM 604145) was found in 21 cases (10.8% - 21/194), corresponding to 26.9% of the group (21/78). In cancer predisposition syndrome conditions, the *BRCA2* gene was the most prevalent, with 19 cases (9.8% - 19/194), corresponding to 26% of this group (19/73); and in miscellaneous group, hereditary amyloidosis associated with the *TTR* gene (MIM 105210) was observed in 37 cases (19% - 37/194), corresponding to 90.2% (37/41).

In the 191 cases with SF, 197 pathogenic/likely pathogenic variants were identified in 33 different genes. The most common genes were *TTR*, with variants in 19% of the cases (37/194), *TTN* in 10.8% (21/194), *BRCA2* in 9.9% (19/194), *LDLR* and *PALB2* in 4.6% each (9/194); *BRCA1* in 4.1% (8/194); *KCNQ1* and *MYBPC3* in 3.6% each (7/194); *PKP2, PMS2* and *TP53* in 3%, each (6/194) (Figure 1). It is worth noting that three cases in our sample had two different secondary findings, so we identified 194 SF in 191 cases. One case had a variant in *LDLR* and a large deletion involving *TMEM127* gene (case 39), the other had variants in *BRCA2* and *TTN* gene simultaneously (case 53) and, the last case had variants in *DSG2* and *PALB2* (case 136) (Table 1 – Supplementary material).

**Figure 1.**
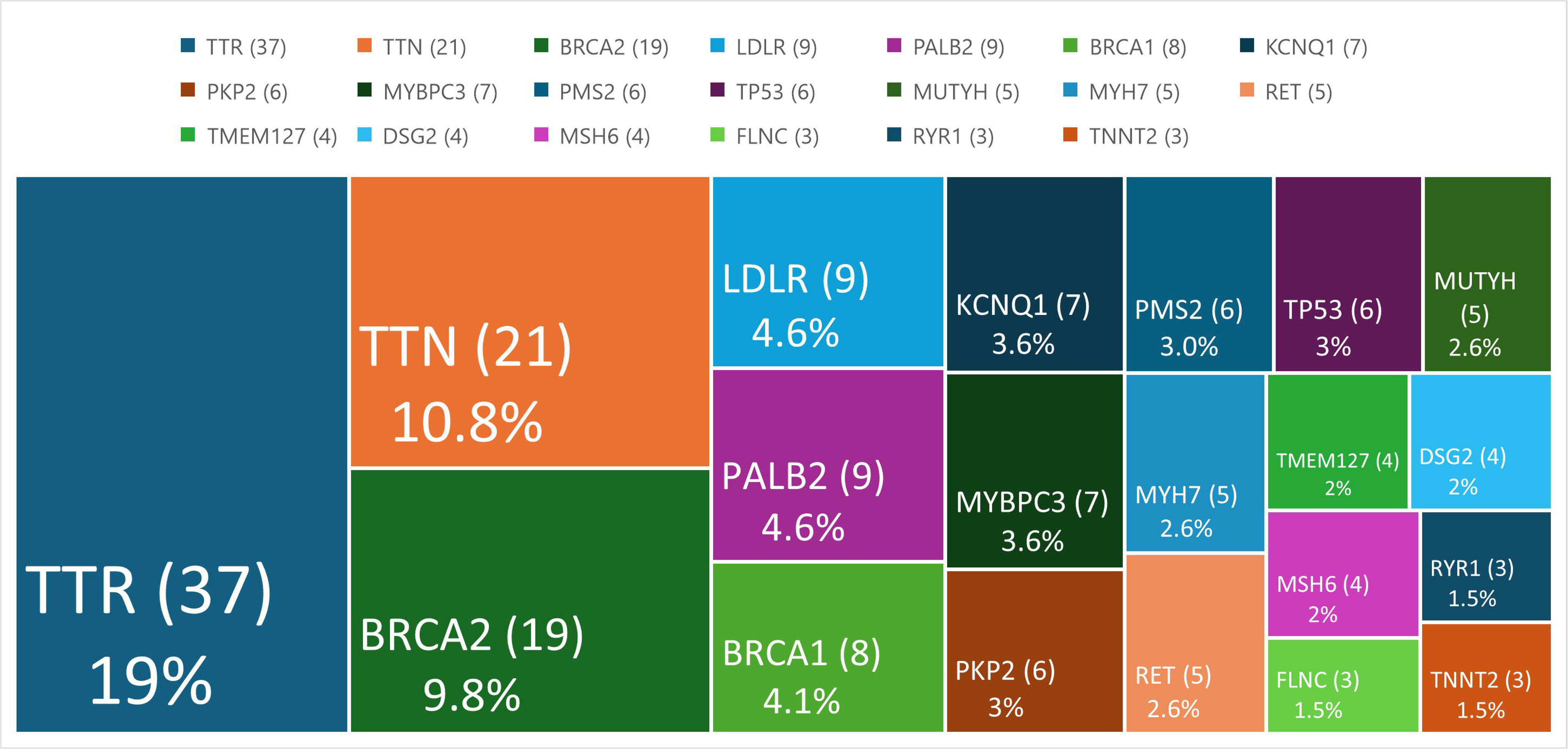
The most frequently identified secondary findings genes (n=194)

We identified 197 P/LP variants (Table 1 – Supplementary material), including 192 single nucleotide variants and small insertions and deletions (SNV/Indels) (97.4% - 192/197) and five structural variants (2.6% - 5/197). The more commonly SNV/Indels encountered were nonsynonymous SNVs (42,7% - 82/192), followed by frameshifts indels (21.3% - 42/192) and nonsense variants (18.2% - 36/192). The SV were all deletions involving only one gene (cases 2, 17, 20 and 173), except for one (case 39). Three of these deletions involved less than two exons of one gene (cases 17, 20 and 173).

Among the 197 identified SF gene variants, several exhibited recurrence across multiple cases, including the *TTR* variant: NM_000371.4:c.424G>A; p.(Val142Ile), found in 33 cases and two variants, which were found in five cases each: the NM_000546.6:c.1010G>A; p.(Arg337His) in *TP53* gene and the NM_000059.4:c.8488-1G>A; p.? in *BRCA2* (Table 1 – Supplementary Material). Additionally, 21 variants (10.6% - 21/197) have not been previously described in the literature and were considered as novel (Table 1 – Supplementary Material).

## DISCUSSION

Literature regarding SF in underrepresented population, through use of genome sequencing, is scarce. The rate of secondary findings in most studies is highly variable, ranging from 0.91% to 8%.^4,17,18,19^ This variability could be attributed to the design, timeframe, population, variant classification guideline used, and genes and variants considered as SF. None of the studies, including ours, demonstrate a gender preference.^4,5,17,18,19^ However, despite this variability, there is a noticeable trend toward a higher rate of these findings over time. This can be attributed, among other factors, to greater access to genomic tests and the incorporation of new genes, as demonstrated by Johnston JJ et al.^20^

Specific studies evaluating SF in Brazilian population, which is large and highly admixed, are scarce.^7, 8^ This gap hinders the assessment of genetic variability across diverse populations and impacts the accurate interpretation of genomic findings, including SF. Although some studies have detailed SF data in the Brazilian population^6,7,8^, to our knowledge, this is the largest study to date, in terms of sample size. These studies, collectively, enhance and broaden the knowledge regarding genomic Brazilian data.

In 2017, Naslavsky et al. described the SF data of 609 healthy elderly Brazilians using the 1.0 version of the ACMG list. They reported a 1.15% rate of SF, including six distinct variants across four genes (*MUTYH*, *BRCA2*, *MSH6,* and *MSH2*), in seven cases.^7^ An interesting finding in this study was the correlation between the genomic finding and the phenotype in elderly individuals, which allowed for a more accurate evaluation of phenotypes that are typically age-related. In two cases there was a personal history of cancer related to the genomic data (*BRCA2* and *MSH6* genes), and in other two, there was none (*BRCA2* gene).^7^ This correlation is often challenging to establish in most studies, as they are generally conducted in younger populations, highlighting the complexity of interpreting and managing such findings.

Naslavsky et al., in 2022, expanding their sample to 1,171 highly admixed healthy elderly Brazilians and, now using the ACMG guideline version 2.0, reported a secondary finding rate of 1.2%. They found 14 different variants among six distinct genes, in 14 cases. The affected genes were *BRCA2, RYR1 -* these two the most frequent - *RET*, *KCNQ1, MYBPC3, SCN5A* and *LDLR.* Only one case was considered as related to the genotype (individual with mild hypercholesterolemia and pathogenic variant in *LDLR* gene).^8^ These two studies by Naslavsky et al. (2017 and 2022) underscore the complexity of these findings, particularly with regard to the access to complete clinical data during the analysis, as they were able to actively verify whether the patient had a personal history related to the SF (In fact, in three cases, the SF could have been considered as a primary in a context outside of a population-based study). However, in most studies and in laboratory practice, the analysis relies on data provided by the healthcare professional, which can be incomplete, making it unlikely to verify whether the phenotype related to SF is present at that specific moment.

Our study showed there was no statistical significance difference in the rate of secondary findings between elderly and non-elderly individuals (< 60 years old versus ≥ 60 years). However, elderly people were underrepresented in our sample due to ascertainment bias, considering that the scope of BRGP is focused on diagnosis of individuals with rare diseases, which often manifest in early life period. We would expect that the rate of SF would be lower in elderly individuals as penetrance for several diseases is age dependent. This could be one of the reasons to explain the lower rate of secondary findings identified in ABraOM sample^7^ when compared to ours (1.2% and 3.6%, respectively).

In 2020, Quaio et al. observed a 7.4% rate of SF in exome sequencing data of 500 individuals with rare diseases. This high rate, however, is justified since they considered as SF not only the 59 genes contained in ACMG-list, but other actionable genes and carrier status for recessive conditions. Considering just SF of ACMG gene list, the rate would decrease to 2.6%.^6^

In our study, the SF rate was 3.6% (191/5,316). We believe that this higher rate when compared to previous studies is because we consistently integrated the ACMG list updates into the analysis within the BRGP and the number of genes in these lists increased significantly. Interestingly, the *TTR* gene was included in the 3.1 version update^16^ (2022) and, coincidently, the most common variant identified in our cohort was *TTR(*NM_000371.4): c.424G>A; p. (Val142Ile) which was found in 33 cases, accounting for 17% of the sample (33/194).

The *TTR* gene was included in the 2022 update based on the justification that individuals of African ancestry are underrepresented in genomic studies, that treatment for the disease is available, and that this inclusion could potentially reduce morbidity. In this way, the addition of *TTR* to the list would improve genomic equity.^16^

This *TTR* variant (c.424G>A; p. (Val142Ile)) is more commonly found in individuals with African ancestry and is associated with a late-onset and not fully penetrant amyloidotic cardiomyopathy.^21^ In 2022, the last Brazilian demographic census showed that 45.3% of the surveyed individuals declared themselves as brown (admixed), 43.5% white, 10.2% black and 0.8% indigenous.^22^ This demographic observation could explain the remarkable *TTR* variant frequency observed in the present study.

Saeidian et al., in 2024, evaluating secondary findings in 16,713 pediatric patients, using v3.2 and beyond ACMG-list, obtained a rate of 5.81% of SF, being *TTR* the most frequently mutated gene, and *TTR*: c.424G>A; p. (Val142Ile), the most identified variant. It is fundamental to highlight that the sample of this study was composed in its majority by African American individuals (51.9% of the sample).^5^ In another large study, conducted by Elfatih et al. (2024), analyzing SF (with non-ACMG as well) in 14,392 individuals from Qatar, the same variant in *TTR* was the most common, supporting the wide prevalence of this variant, including in Arabic population.^3^

Therefore, considering that the variant c.424G>A; p.(Val142Ile) has a high prevalence in people of African ancestry and, consequently, among Brazilians, and that the phenotype associated to this variant is late-onset with incomplete penetrance, we raise concerns regarding the psychological effects to the individuals and the economic burden that it can cause, considering that many individuals will have this variant disclosed, and consequently should be screened for cardiomyopathy, even though many of these patients will not develop any disease.

Besides *TTR,* the most identified genes reported in studies concerning SF were *TTN, RYR1,* and *BRCA2.*^3,4,5,18,19,23^. In our study we observed that *TTN* and *BRCA2* were genes also commonly reported (Figure 1). These findings suggest a potential pattern in terms of genes more frequently observed as SF, even across different populations. However, acknowledging distinct findings offers greater benefit, as they may be unique to a specific population. In addition to the variant previously discussed in *TTR*, two other variants were recurrent in our study, highlighting these genomic peculiarities.

The *TP53* variant NM_000546.6: c.1010G>A; p. (Arg337His), known as R337H, is a variant commonly identified in this gene in Brazilians from South and Southeast region, due to a founder effect.^24^ Not surprisingly, it was the second most common variant detected as SF in our study (cases 55, 58, 61, 165, 169), with all cases originating from participant centers in these two regions. This gene is associated with Li-Fraumeni syndrome (LFS) (MIM 151623), a hereditary cancer predisposition syndrome that has high penetrance and a high cumulative risk for the development of various types of cancer, including multiple tumors, even in childhood.^24,25^ Similar to other cancer phenotypes included in the ACMG guidelines, LFS has an established tumor screening protocol that can be initiated at any age. In our cohort, three cases were individuals under 18 years of age. These screening protocols are associated with reduced morbidity and mortality and improved survival outcomes, primarily due to early detection, which can result in greater cost-effectiveness.^25^ However, particularly within the context of Brazil’s public healthcare system, access to these screening tests remains limited, highlighting concerns about the system’s capacity to adequately manage individuals with rare diseases or carriers, including not only cancer-related phenotypes but also other conditions. As access to genomic testing continues to expand and SF becomes more commonly reported, it is essential for the healthcare system to be adequately prepared to address these emerging needs, otherwise the actionability for the ACMG-genes will only be theoretical. In other words, to be a truly actionable gene, once the SF is identified in an individual, the access to adequate treatment, screening protocols, genetic counseling, including familial genetic testing, should be universally guaranteed by the public healthcare system. Unfortunately, this is not the scenario in many countries, including Brazil.

Although for LFS there is an established tumor screening protocol, including the childhood period, for some genes, such as *BRCA1* and *BRCA2,* there is no specific tumor screening protocol to be established in childhood to reduce morbimortality. Indeed, *BRCA1* and *BRCA2-linked* tumors are expected to manifest in adulthood. ^26^ In our sample, 63.9% of the secondary findings were identified in individuals < 18 years old. The team of BRGP decided to disclose ACMG gene list of SF regardless of individual’s age, mainly considering an expanded benefit for its family. However, we are aware of ethical issues related to reports of specific SF in children, as they may not represent a true benefit for the children when they are disclosed. In 2021, Hunter et al., addressing the actionability of SFs in children and adolescents, established the Pediatric Actionability Working Group (PAWG) and revisited the list of actionable SF in children and adolescents. ^26^ Of the 81 genes on the current ACMG list ^2^, 46 are classified as having definitive or strong actionability by the PAWG. Among the 122 individuals in our cohort under 18 years of age, 33.6% (41/122) had SF involving these 46 genes. Regardless of the SF reporting policy, we believe that pre-test genetic counseling is essential, as it ensure that individuals are fully informed about the potential outcomes and enables them to make informed decisions about whether to accept or decline these findings.

The *BRCA2* variant NM_000059.4: c.8488-1G>A, which was also identified in five of our cases (cases 26, 60, 119, 142, 161), is a known pathogenic variant, described in different populations. It has been already identified in Brazilian patients, mainly in individuals with Portuguese heritage.^27^ Das Virgens CS, in 2023, analyzing data of individuals from Bahia - a Northeast region state in Brazil - with P/LP variants in *BRCA1* and *BRCA2* genes - observed that the c.8488-1G>A was the most frequently identified *BRCA2* variant.^27^ Other studies with Brazilian samples, including our own, corroborate that this variant It is one of the most common *BRCA2* variants in Brazil.^28,29,30^

Among the 197 variants observed in our study, the majority were SNV/indels as seen in other studies.^3,5,18,31^ However, we identified five deletions. All involved a single gene, except one, a 470.5 kbp deletion that involved ten protein coding genes, including the haploinsufficient gene *TMEM127*, associated with susceptibility to pheochromocytoma (MIM 171300). Recommendations related to SF are primarily made concerning genomic sequencing findings, particularly involving SNVs/Indels and small SV^32,33^. However, regardless of the test performed, whether sequencing or chromosomal microarray (CMA), copy number variants (CNV) can be detected, potentially involving SF genes. In many SF studies, structural variants are frequently excluded from the data, even when GS is employed.^3,5,18^ This data is less discussed in the literature, and the few studies that address them report a rate of SF of 0.26% to 0.29% in CMA results.^32,33^ These lower rates can mainly be attributed to the fact that most of these findings may be considered as a primary finding, as they involve more than one gene. Nevertheless, CNV analysis should not be overlooked, especially with the increased availability of GS and exome sequencing (ES), which also allows CNV detection.^32,33^ Moreover, three of the SV observed in our study involved less than two exons (cases 17, 20 and 173). We believe that these variants could be missed by ES.

We identified 21 novel variants (Table 1 – Supplementary Material). These unique variants also underscore the importance of genomic diversity among populations, as many of these variants may be specifically associated with certain populations.^19^

In our study we identified three cases (cases 39, 53 and 136) that carried two SF in different genes (Table 1 – Supplementary material), accounting for 1% of the cases, which is in accordance with other studies.^4,19,23^

Regarding the major phenotypic groups of SF, the most prevalent in the literature are cardiovascular conditions, cancer predisposition syndromes and miscellaneous.^3,4,5,18,19,23^ Inborn errors of metabolism phenotypes are the less represented, even in studies with high rates of consanguinity^19^, which may be explained by the smaller number of involved genes (four in the ACMG 3.2 version 2023 update) and the inheritance pattern of this group of diseases is usually autosomal recessive.^5^ Our study also supports these data (Figure 2).

**Figure 2.**
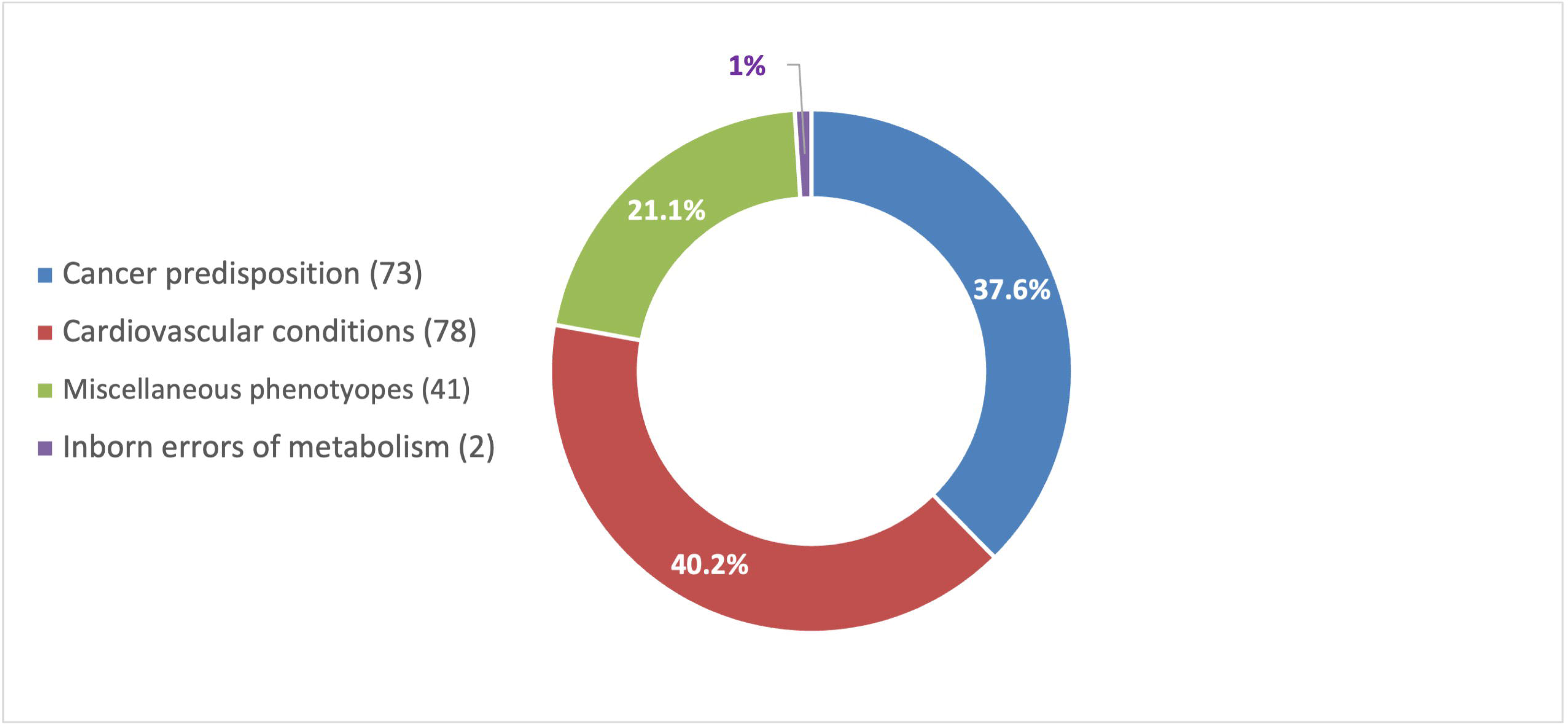
Frequency of the different groups of conditions related to SF (n=194)

In summary, our study described the profile of SF in 5,316 individuals of BRGP. The rate and profile of genomic secondary findings vary across populations. In our study, we identified recurrent and novel variants in a Brazilian population with rare diseases, with *TTR, TTN* and *BRCA2* being the most affected genes. The most identified SF phenotypes were cardiovascular conditions, cancer predisposition syndromes and miscellaneous. While SF are intended to bring benefits as discovered, some of these findings remain debatable regarding their selection, as they may pose a burden to the patient, their family, and to the healthcare system. Our data also highlight the need for public policies and access to genetic counseling for better management of individuals with rare diseases and secondary findings.

## Data Avaliability

The anonymized data supporting the findings of this study are provided in Table 1 in supplementary material. Further details regarding the data and variants can be obtained upon request from the corresponding author.

## Acknowledgments

We would like to express our gratitude to the participants of this study and their families. This research was made possible by the data and findings provided by the Rare Genomes Project, an initiative of the Hospital Israelita Albert Einstein (HIAE) in collaboration with the Programa de Apoio ao Desenvolvimento Institucional do Sistema Único de Saúde (PROADI-SUS) of the Brazilian Ministry of Health (Law 12.101/2009). We also extend our thanks to the reference participant centers involved in this project.

## Authors Contribution

Conceptualization: E.P., L.V.; Data curation: E.P., L.V; Formal analysis: E.P., L.V; Methodology: E.P., L.V.; Resources: E.P., L.V.; A.V.C.C; Writing-original draft: E.P., L.V; Visualization, Writing-review & editing: E.P.; L.V.; A.V.C.C.; M.F.; C.A.M.; J.R.M.P.; J.G.A.E.; M.M.; T.Y.T.S.; C.R.D.C.Q.; J.R.M.C.; K.C.; R.M.M.; A.C.B.T.; R.Y.Y.; V.P.C.; L.S.S.; K.O.P.; J.B.O.F.; and T.F.A.

## Ethical Declaration

The BRGP and related studies were submitted to the ethics committees of HIAE and received approval (CAAE: 9567220.4.1001.0071.).

An informed consent was obtained from each participant, which includes the option to accept or decline receiving results related to secondary findings (following recommendations by ACMG). The written patient’s consent was received and archived. The genomic data of participants were de-identified.

